# Association of Mass Gatherings and COVID-19 Hospitalization

**DOI:** 10.1101/2020.10.27.20220707

**Authors:** Oren Miron, Kun-Hsing Yu, Rachel Wilf-Miron, Nadav Davidovitch

**Affiliations:** Ben-Gurion University, Beer Sheva, Israel; Harvard Medical School, Boston, Massachusetts; Tel Aviv University, Tel Aviv, Israel

**Keywords:** COVID-19, SARS-CoV-2, Mass Gathering, Hospitalization

## Abstract

We examined COVID-19 hospitalizations following mass gatherings in Wisconsin and Minnesota, United States (September 17-18, 2020). We found that the hospitalization rate increased 15-fold in the Minnesota gathering county, and 12.7-fold in the Wisconsin gathering county. On the state level, it increased 2-fold in Minnesota, and 2.3-fold in Wisconsin, while not increasing significantly in states without gatherings. Our findings suggest that mass gatherings are followed by increased COVID-19 hospitalizations, and that precautions should be taken.

## Introduction

In march 2020, at the start of the Coronavirus Disease-2019 (COVID-19) pandemic, most countries and organizations recommended to avoid mass gatherings.[1] In September 2020, there have been policy makers that brought back mass gatherings under the assumption that it can now be done safely.

One such gathering was in an outdoor rally in Minnesota, United States, which was followed by COVID-19 hospitalization of two of its participants.[2] It is not known if they were infected in the gathering, and if the rate of hospitalizations changes following mass gatherings.[3,4]

## Methods

We extracted COVID-19 hospitalization by date at the county and state level for the mass gatherings in outdoor rallies in Beltrami, Minnesota (gathering on September 18, 2020), and Marathon, Wisconsin (gathering on September 17, 2020).[5]

Those counties had a combined population of 182,880 residents based on the U.S. Census data. We analyzed the county incidence rate per 100,000 capita and utilized a weekly moving average. We compared the change from the gathering day to 21 days later, since that was the most days of data we had on both counties, and since most infections reach hospitalizations during that period.[6]

We also examined the change in the relevant states of the gathering counties (Wisconsin, Minnesota). We compared that change with the change in states who did not have a mass gathering rally since April 2020 and had daily hospitalizations counts in COVID-tracking project.

These states were Alabama, Arkansas, Colorado, Connecticut, Georgia, Hawaii, Idaho, Indiana, Kansas, Kentucky, Massachusetts, Maryland, Maine, Mississippi, Montana, North Dakota, Nebraska, New Jersey, New Mexico, New York, Oklahoma, Oregon, Rhode Island, South Carolina, South Dakota, Tennessee, Utah, Washington, and Wyoming. For non-gathering states, we used a comparison date that was the date of the Minnesota gathering (September 18, 2020). Statistical significance was evaluated based on the 95% confidence interval.

## Results

In Beltrami, on the gathering day the hospitalization rate was 0.3/100,000 (95% confidence interval -0.2 to 0.8). By day 21 the hospitalization rate increased to 4.5/100,000 (95% confidence interval 1.5 to 7.6; 15-fold).

In Marathon, on the gathering day the hospitalization rate was 0.3/100,000 (95% confidence interval 0 to 0.6). By day 21 the hospitalization rate increased to 3.8/100,000 (95% confidence interval 2.5 to 5.1; 12.7-fold; figure 1).

**Figure 1:**
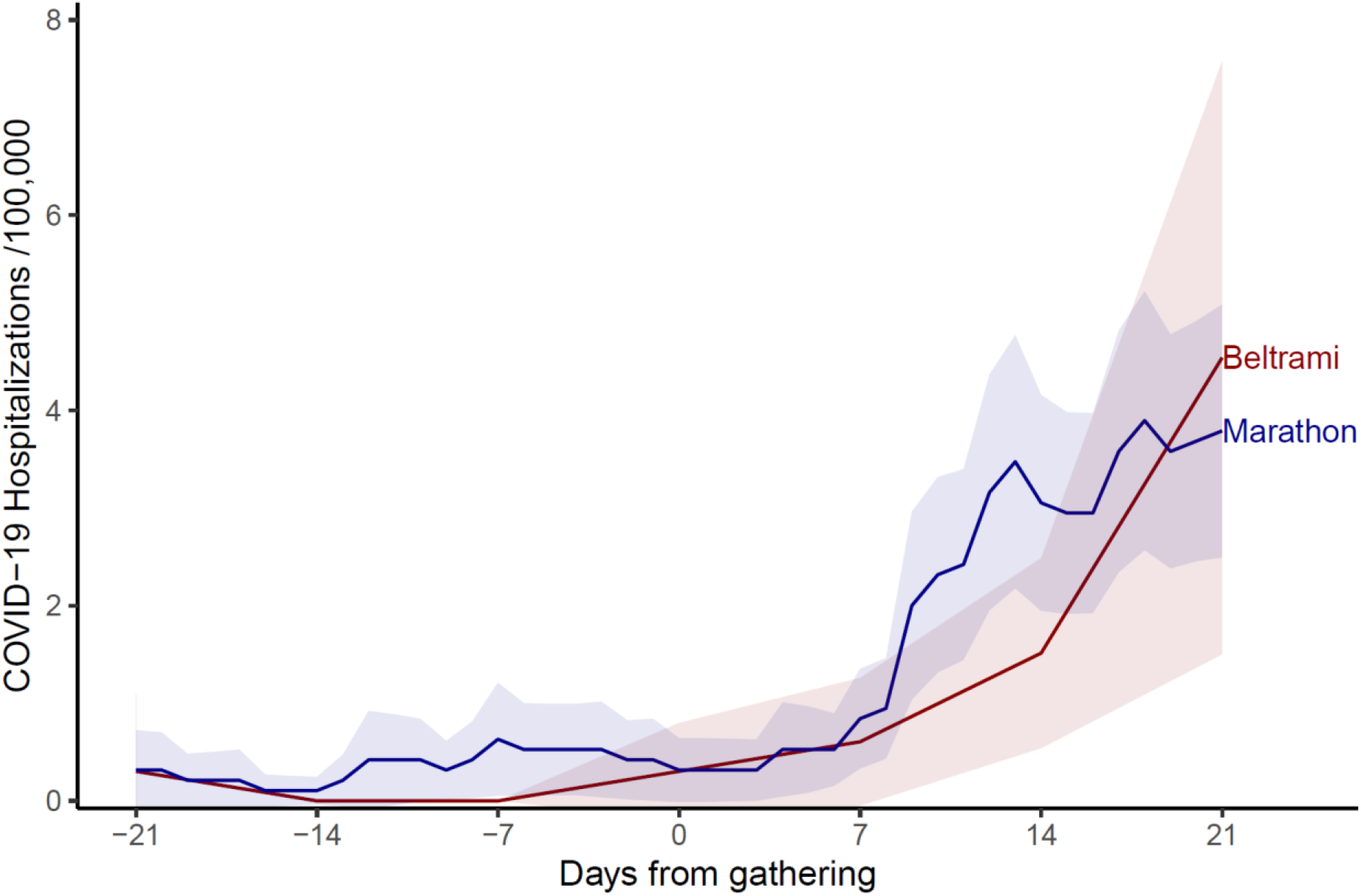
COVID-19 hospitalizations in mass gathering counties. Legend: Coronavirus Disease-19 (COVID-19) hospitalizations by days from gathering. Y-axis indicates COVID-19 daily hospitalizations per 100,000 capita after a 7-day moving average. X-axis indicates the date. The red line indicates Beltrami county, Minnesota, and the blue line indicates Marathon county, Wisconsin. The colored area indicates the 95% confidence interval. Marathon county reported hospitalizations daily, while Beltrami county reported it weekly on Fridays, which we used to calculate its daily rate.

In Minnesota, on the gathering day the hospitalization rate was 0.6/100,000 (95% confidence interval 0.5 to 0.7). By day 21 the hospitalization rate increased to 1.2/100,000 (95% confidence interval 1.1 to 1.3; 2-fold; figure 2).

**Figure 2:**
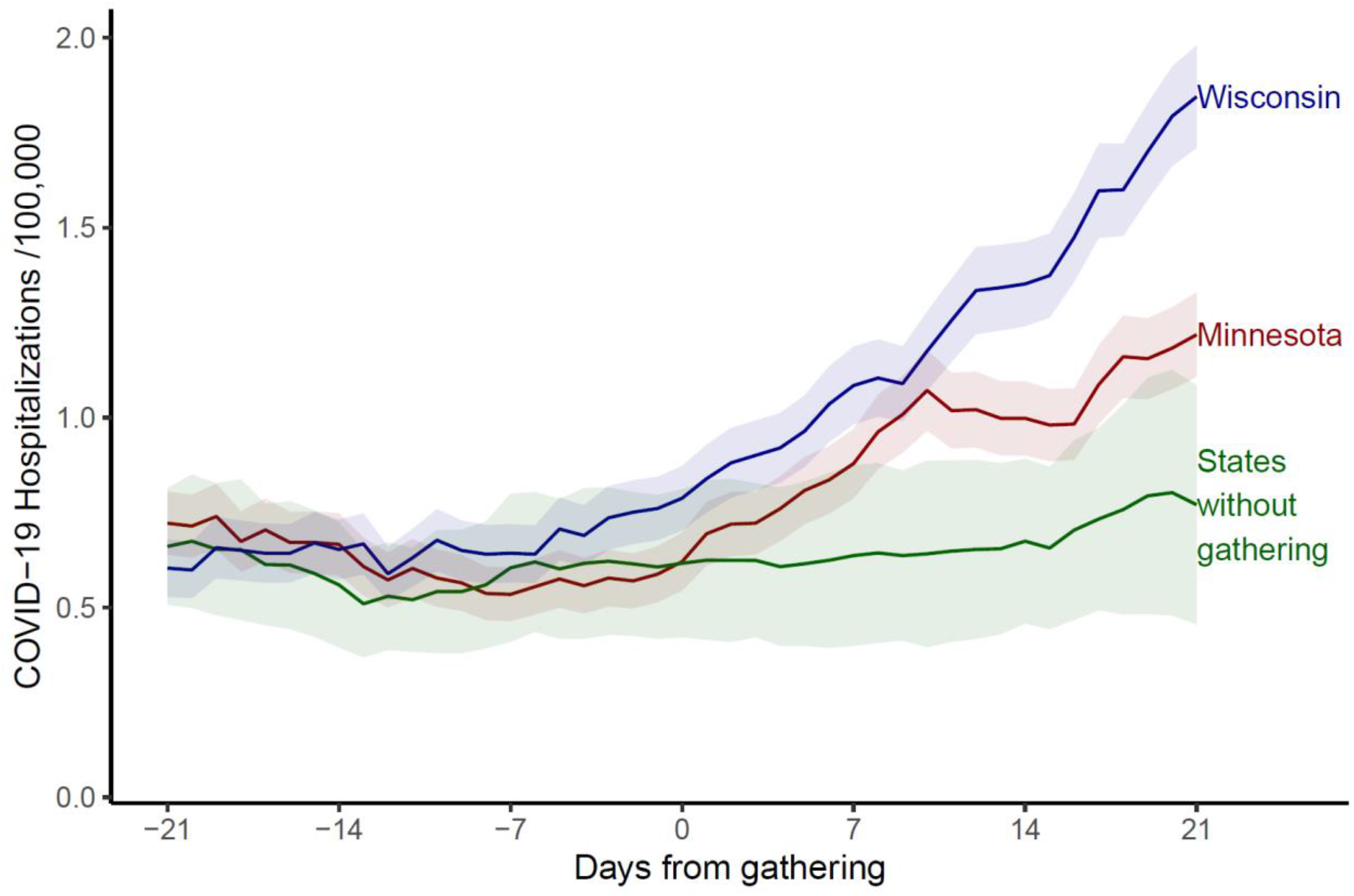
COVID-19 hospitalizations in mass gathering states. Legend: Coronavirus Disease-19 (COVID-19) hospitalizations by days from gathering. Y-axis indicates COVID-19 daily hospitalizations per 100,000 capita after a 7-day moving average. X-axis indicates the date. The red line indicates Minnesota, the blue line indicates Wisconsin, and the green line – other U.S. states without similar mass gatherings.

In Wisconsin, on the gathering day the hospitalization rate was 0.8/100,000 (95% confidence interval 0.7 to 0.9). By day 21 the hospitalization rate increased to 1.8/100,000 (95% confidence interval 1.7 to 2; 2.3-fold; figure 2).

In states without mass gatherings, on the comparison day the hospitalization rate was 0.6/100,000 (95% confidence interval 0.4 to 0.8). By day 21 the hospitalization rate increased to 0.8/100,000 (95% confidence interval 0.5 to 1.1; 1.3-fold; figure 2).

## Discussion

Our analysis showed that mass gatherings were followed by an increase in COVID-19 hospitalizations on the county and state levels. The gatherings occurred in late September, and during this period the hospitalizations in states without a mass gathering did not increase significantly. Our main limitation is that both Wisconsin and Minnesota had mass gatherings a month before the analyzed gatherings, which could have caused a second wave of infection that would affect their hospitalizations a month later. Minnesota also had a mass gathering two weeks after the analyzed mass gatherings, which could explain why its rate started rising again in early October.

Our findings suggest that precautions such as masks, should be taken in mass gatherings such as political rallies, large school assemblies and cultural events, to reduce COVID-19 hospitalizations.[7] Since these precautions are often not taken by participants of the gatherings, there is also a need for the state’s policy makers to increase public health measures such as social distancing and face masks wearing in the geographic area close to where the rally takes place.[8] Also, it is advised to prepare the healthcare infrastructures (including the hospitals) to prepare for the possible increase in COVID-19 hospitalizations.[9]

## Supporting information

STROBE checklist

## Data Availability

Data availability statement: data is publicly available at the state level at https://covidtracking.com/data/download and at the county level for Beltrami https://covid-19-response-beltramicounty.hub.arcgis.com and Marathon https://data.dhsgis.wi.gov/datasets/covid-19-historical-data-by-county 

https://covid-19-response-beltramicounty.hub.arcgis.com

https://data.dhsgis.wi.gov/datasets/covid-19-historical-data-by-county

## Author contribution

Concept and design: All authors.

Acquisition, analysis, or interpretation of data: All authors. Drafting of the manuscript: Oren Miron.

Critical revision of the manuscript for important intellectual content: Yu, Wilf-Miron, and Davidovitch.

Statistical analysis: Oren Miron and Yu.

Study supervision: Davidovitch.

